# Dysregulation of lncRNA MALAT1 Contributes to Lung Cancer in African Americans by modulating the tumor immune microenvironment

**DOI:** 10.1101/2024.04.04.24305363

**Authors:** Jin Li, Pushpa Dhilipkannah, Van K Holden, Ashutosh Sachdeva, Nevins W Todd, Feng Jiang

## Abstract

African American (AA) populations present with notably higher incidence and mortality rates from lung cancer in comparison to other racial groups. Here, we elucidate the contribution of long non-coding RNAs (lncRNAs) in the racial disparities and their potential clinical applications in both diagnosis and therapeutic strategies. AA patients had elevated plasma levels of MALAT1 and PVT1 compared with cancer-free smokers. Incorporating these lncRNAs as plasma biomarkers, along with smoking history, achieved 81% accuracy in diagnosis of lung cancer in AA patients. We observed a rise in MALAT1 expression, correlating with increased levels of monocyte chemoattractant protein-1 (MCP-1) and CD68, CD163, CD206, indicative of tumor-associated macrophages in lung tumors of AA patients. Forced MALAT1 expression led to enhanced growth and invasiveness of lung cancer cells, both in vitro and in vivo, accompanied by elevated levels of MCP-1, CD68, CD163, CD206, and KI67. Mechanistically, MALAT1 acted as a competing endogenous RNA to directly interact with miR-206, subsequently affecting MCP-1 expression and macrophage activity, and enhanced the tumorigenesis. Targeting MALAT1 significantly reduced tumor sizes in animal models. Therefore, dysregulated MALAT1 contributes to lung cancer disparities in AAs by modulating the tumor immune microenvironment through its interaction with miR-206, thereby presenting novel diagnostic and therapeutic targets.

## INTRODUCTION

Lung cancer stands as the foremost cause of cancer-related fatalities among both men and women in the USA(1). Non-small cell lung cancer (NSCLC) constitutes 85% of all lung cancer cases and primarily comprises two histological types: adenocarcinoma (AC) and squamous cell carcinoma (SCC) (1). Significant disparities in NSCLC exist across various ethnicities, with African Americans (AAs) facing a higher incidence and mortality rate from this disease (2). Apart from socioeconomic disparities, biological factors such as tumor biology, genetics, and molecular changes also contribute to disparities in lung cancer (3). Genome-wide association studies have pinpointed specific regions on chromosomes 5p15 and 15q25 as lung cancer susceptibility loci within the AA population(4). Differences in the methylation levels of genes with functional significance, such as the nuclear receptor subfamily 3, have been identified as potential contributors to the racial disparities observed in NSCLC(5). Furthermore, the epidermal growth factor receptor mutation is more prevalent in AA lung cancer patients compared to other populations (6). Additionally, heightened levels of cytokines, including IL-1β, IL-10, and TNFα, have been linked to an increased risk of lung cancer within the AA population (7, 8). However, the mechanisms underlying lung cancer disparities in AA remain unclear.

Long non-coding RNAs (lncRNAs), longer than 200 nucleotides in length, play pivotal roles in various aspects of cancer development, progression, and metastasis(9). LncRNAs are a crucial regulatory layer in lung cancer, playing diverse roles in driving the tumorigenesis (9). For instance, lncRNAs modulate key signaling pathways, including PI3K/AKT and MAPK, impact metastasis of NSCLC through epithelial-mesenchymal transition, and influence immune responses(10). MALAT1 (metastasis associated lung adenocarcinoma transcript 1) is associated with metastatic potential of NSCLC, while TUG1 can affect immune checkpoint molecules, potentially influencing immunotherapy responses(11, 12). Previous research, including our own, has identified lncRNAs linked to lung cancer, underscoring their promise as potential biomarkers for this disease (13, 14). However, the earlier studies on lncRNAs have predominantly focused on individuals of White Ancestry. We postulate that investigating the roles of lncRNAs in cancer biology, including their influence on tumorigenesis and immune regulation within the AA population, holds the potential to ameliorate racial disparities in lung cancer by enhancing diagnostics and therapeutics.

## MATERIALS AND METHODS

### Patients and specimens

Our study was approved by the Institutional Review Board of the University of Maryland Baltimore. We recruited lung cancer patients and cancer-free smokers based on specific criteria complemented by demographic, radiological, and clinical data from their medical records (1) . Blood samples were collected from the participants and plasma was separated from blood samples within one hour of collection by following established clinical protocols (13, 15, 16). 92 NSCLC patients and 76 cancer-free smokers were enrolled, including 38 AA and 54 WA lung cancer patients, along with 35 AA and 41 WA cancer-free smokers (Table 1). Among 92 NSCLC patients, stages I to IV were represented, with 52 cases of adenocarcinoma and 40 of squamous cell carcinoma. The 48 cancer-free smokers presented with various benign conditions. Furthermore, surgically resected lung tumor tissues and their adjacent non-cancerous counterparts were collected from 46 NSCLC patients, including 23 AAs and 23 WAs. These patients underwent lobectomy or pneumonectomy. Detailed demographics and clinical characteristics are in Table 1.

**Table 1.** Characteristics of NSCLC patients and cancer-free smokers.

| Table 1. Characteristics of NSCLC patients and cancer-free smokers |  |  |  |
| --- | --- | --- | --- |
| Plasma specimens from patients with NSCLC and cancer-free smokers were analyzed. |  |  | Tissue specimens were analyzed from forty-six patients with NSCLC |
|  | NSCLC cases (n = 92) | Controls (n = 76) |  |
| Age | 69.13 (SD 11.37) | 67.85 (SD 10.34) | 68.3 ± 7.5 |
| Sex |  |  |  |
| Female | 24 | 20 | 29 |
| Male | 68 | 56 | 17 |
| Race |  |  |  |
| African Americans | 38 | 35 | 23 |
| White Americans | 54 | 41 | 23 |
| Smoking pack-years (median) | 32.9 | 26.5 | 39 |
| Stage |  |  |  |
| Stage I | 23 |  | 24 |
| Stage II | 35 |  | 22 |
| Stage III-IV | 34 |  | 8 |
| Histological type |  |  |  |
| Adenocarcinoma | 52 |  | 26 |
| Squamous cell carcinoma | 40 |  | 20 |

Abbreviations: NSCLC, non-small cell lung cancer. SD, standard deviation.

### Droplet digital PCR (ddPCR)

43 lung cancer-associated lncRNAs were selected from the prior studies (Supplementary Table 1)(14, 17, 18). ddPCR was conducted to measure the copy numbers of the 43 lncRNAs following the method previously outlined(18). Briefly, RNA was first reverse transcribed using the TaqMan miRNA RT Kit. The ddPCR reactions, comprising a mix of cDNA, Supermix, and Taqman primer/probe mix, were processed in the QX100 Droplet Generator (Bio-Rad Laboratories, Hercules, CA), generating over 10,000 droplets per well. Post-PCR amplification, these droplets were examined using a fluorescence detector to accurately detect lncRNA levels. We used Poisson’s distribution to determine the concentration of target genes, ensuring precise quantification.

### Cell culture

We acquired six human NSCLC cell lines (A549, H513, H1373, H1385, H460, and H520) and the normal lung cell line (BEAS-2B) from the American Type Culture Collection (ATCC, Manassas, VA). H513, H1373, and H1385 are NSCLC cell lines originally developed from AA lung cancer patients, while A549, H460, and H520 were derived from WA lung cancer patients. These cell lines were cultured according to the manufacturer’s instructions. The culture medium was carefully removed and centrifuged at 3,000 revolutions per minute for 10 minutes at 4°C to collect the supernatants, which contained the secreted factors from the cells, free of cellular debris or intact cells.

### Cell transfection

To overexpress MALAT1, we amplified the full-length human MALAT1 cDNA and cloned it into the pcDNA3.1(+) mammalian expression vector (Invitrogen, Carlsbad, CA) using KpnI and XhoI restriction sites, resulting in the construct pcDNA3.1-MALAT1(19). The full-length human MALAT1 cDNA was amplified using the following primers: forward, 5’-GGCGGTACC ATGAAACAATTTGGAGAAG -3’; reverse, 5’-GCGCTCGAGCTAAGTTTGTA-CATTTTGCC -3’. Stable cell lines overexpressing MALAT1 were established by selecting for G418-resistant colonies. For control experiments, we utilized the empty pcDNA3.1(+) vector (pcDNA3.1-NC). For MALAT1 knockdown, MALAT1-targeting short hairpin RNA (shRNA) (5’-TCCACTT-GATCCCAACTCATC-3’; MALAT1-shRNA2, 5’-TTCCTTAGTTGGCATCAAGGC-3’) and a non-targeting scrambled shRNA control (5’-GACCTGTACGC-CAACACAGTG-3’), synthesized by Integrated DNA Technologies (IDT, Coralville, IA), were cloned into the psiSilencer 4.1-CMV neo vector (Invitrogen), with puromycin selection employed for stable integration(19). Plasmids were prepared using the Qiagen DNA Midiprep Kit (Qiagen, Santa Clarita, CA). To either enhance or suppress miR-206 expression, we utilized synthesized miR-206 mimic (GGUGUGUGAAGGAAUGUAAGGU), NC-mimic (GCGUCUCGAUGCCGAGAGCUA), antisense oligoribonucleotides (ASO, ACCUUACAUUCCUUCACACACC), and NC-ASO (TAGCUCUCGGCAUCGAGACGC), respectively, obtained from IDT(20). Transfection into cancer cell lines was performed using Lipofectamine 2000 (Life Technologies, Gaithersburg, MD) for experiments involving MALAT1, and Lipofectamine RNAiMAX (Life Technologies) for those involving miR-206.

### MTT Assay (3-(4,5-Dimethylthiazol-2-yl)-2,5-diphenyltetrazolium bromide)

MTT assay was utilized to evaluate cell proliferation, as outlined in prior studies(21, 22). Cells were seeded at a density of 5,000 cells per well in a 96-well plate and left to adhere overnight in a 37°C incubator with 5% CO_2_. Subsequently, 100 μL of fresh medium containing MTT at a final concentration of 0.5 mg/mL was added to the wells and the cells were incubated at 37°C. After the incubation period, the MTT solution was removed and 100 μL of Dimethyl sulfoxide (Sigma-Aldrich) was added to each well to dissolve the formazan crystals. The absorbance at 570 nm was then measured using a microplate reader. To assess cell viability, the average absorbance of each treatment group was calculated and normalized against the control group.

### Transwell assays

Cell migration was determined by using the Transwell assay (AMSBIO LLC, Cambridge, MA) (21, 22). The cells that had migrated to the lower surface of the membrane were fixed with formalin and stained with crystal violet (Sigma-Aldrich). The migrating cells were examined microscopically and determined by counting the migrating/invasive cells in 5 randomly selected fields using an Olympus BX41 microscope (Olympus Corporation).

### Matrigel invasion assay

5 × 10^4^ cells are seeded in the upper chamber of a Transwell apparatus (Corning Inc., Corning, NY), coated with Matrigel (BD Biosciences, Franklin Lakes, NJ). The lower chamber contains serum. After incubation, cells that invade through the Matrigel and reach the lower chamber are fixed with methanol, stained with crystal violet, and counted to assess their invasive capability.

### Dual luciferase reporter assay

To investigate MALAT1 and miR-206 interaction, MALAT1 sequence containing miR-206 binding sites was inserted into a pmirGLO dual luciferase vector (Promega, Madison, WI) to generate wildtype (WT) pmirGLO-MALAT1. The mutant (MUT) of MALAT1 sequence in miR-206 binding sites was synthesized using Site-Directed Mutagenesis Kit (Thermo Fisher Scientific) and inserted into a vector to generate MUT pmirGLO-MALAT1. Custom miR-206 mimics and inhibitors were purchased from Horizon Discovery (Boyertown, PA). The pmirGLO vectors containing WT-, or MUT-MALAT1 sequence were co-transfected with miR-206 mimic and miR-206 inhibitor to cancer cells by Lipofectamine2000 (Invitrogen, Carlsbad, CA). The relative luciferase activities were measured by Dual-Luciferase Reporter Assay protocol (Promega). To investigate miR-206 and MCP-1, the 3′UTRs of MCP-1 containing the predicted miR-206-binding sites and MUT sites were respectively inserted into pmirGLO dual luciferase vector pmirGLO-MCP-1-3′UTR-WT and pmirGLO-MCP-1-3′UTR-MUT. The 3′UTRs of MCP-1 with predicted miR-206-binding sites and their mutant versions were cloned into pmirGLO vectors. These constructs were co-transfected into lung cancer cells with miR-206 mimic and inhibitor. The same Dual-Luciferase Reporter Assay was used to evaluate the regulatory effect of miR-206 on MCP-1.

### RNA pull-down assay

H292 cells (3 × 10^6^) were cultured on 10-cm plates for 24 hours and transfected with 100 nM of either biotin-labeled WT-miR-206, biotin-labeled MUT-miR-206, or biotin-labeled negative control (NC). Cells were harvested 48 hours post-transfection. Streptavidin-coated Dyna beads (Invitrogen) were prepared by coating with 10 μL yeast tRNA (Thermo Fisher Scientific) and incubated in lysis buffer. After washing, 600 μL of cell lysates were mixed with 50 μL of pre-coated beads and incubated for 4 hours. Beads were pelleted to remove unbound material at 4°C for 2 minutes at 500g. MALAT1 levels in the pulldown samples were quantified using ddPCR.

### Western blot analysis

Protein samples (30 µg) were separated via Sodium Dodecyl Sulfate Polyacrylamide Gel Electrophoresis and transferred to Polyvinylidene Difluoride membranes (MilliporeSigma, Saint Louis, MI). The membranes were first subjected to blocking to prevent non-specific binding and then incubated with primary antibodies specifically targeting the proteins of interest (Abcam, Waltham, MA). Secondary antibody incubation was followed by detection using an enhanced chemiluminescence reagent (MilliporeSigma).

### The Luminex® multiplex assays

The Luminex® Multiplex Assays (Thermo Fisher Scientific) was used for the quantification of proteins in plasma based on the manufacturer’s instructions. Tissue and cellular samples were lysed and subsequently processed according to the assay’s protocol(23). The plasma samples, along with the prepared tissue and cellular lysates, were then incubated with specifically color-coded beads. Each bead set was coated with antibodies targeting one of the aforementioned proteins, enabling the simultaneous detection and quantification of these key biomarkers in our samples.

### Human lung cancer subcutaneous xenografts in mice

With an animal study protocol approved by the University of Maryland Baltimore, we utilized six-week-old female immunodeficient NOD. Athymic Swiss nu/nu/Ncr nu (nu/nu) mice (The Jackson Laboratory, Bar Harbor, ME). To assess the in vivo oncogenic function of MALAT1. The mice were randomly divided into two groups (n=5/group). One group was injected subcutaneously with 1 × 10^6^ NCI-H460-luc cells (TD2 Precision Oncology, Scottsdale, AZ) transfected with pcDNA3.1-MALAT1. Another group, the control group, received an equal number of the cancer cells transfected with the empty pcDNA3.1(+) vector (pcDNA3.1-nc). Tumor volumes were monitored on a weekly basis using In Vivo Imaging System (IVIS) 200 Imaging System (Xenogen, Alameda, CA) following a previously described method(22). After four weeks, the mice were euthanized under deep anesthesia using pentobarbital (Sigma-Aldrich), and Tumor sizes were determined using the formula for tumor volume: Tumor volume = 1/2(length × width2). Tissue samples were harvested for further analysis.

To assess the efficacy of MALAT1 targeting, twenty mice were subcutaneously injected with A549-Luc cells (CCL-185-LUC2™, ATCC) at a density of 5 × 10^5^ cells in 50 μl of RPMI medium. This mixture was thoroughly combined with an equal volume of Corning® Matrigel matrix (Corning) before being subcutaneously inoculated into each mouse’s back. Seven days later, marking the treatment’s commencement (day 0), the mice were segregated into two groups: one group received a control ASO treatment, while the other was administered a MALAT1-targeting ASO (AstraZeneca, Wilmington, DE). Each group received their respective treatments subcutaneously at a dose of 50 mg/kg, administered weekly for three weeks. Tumor volumes were monitored on a weekly basis using IVIS. Three weeks after treatment, five mice from each group were euthanized, and their tumor sizes were documented. Tissue samples were collected for additional analysis. The remaining mice continued to be observed until their natural demise.

### Immunohistochemical analysis of tumor tissues

Tissues were fixed, paraffin-embedded, and sectioned. Sections underwent antigen retrieval, incubation with primary antibodies (Abcam), and secondary antibody treatment. Chromogen development and counterstaining were performed, followed by microscopy (Olympus Corporation). For the quantitative analysis, five representative regions encompassing both cell and stroma areas were carefully selected. The positive cells were counted, and an average was calculated for each area. The resulting values for every hundred cells were classified according to the following scale: 0 (0%-5% positive cells), 1 (6%-25% positive cells), 2 (26%-50% positive cells), and 3 (more than 50% positive cells)(24). Cells marked by a distinct, strong brown stain were manually counted using the AnalySIS Pro software (Olympus). The counting process was independently performed three times for each area by two scientists, ensuring objectivity. All image analyses were conducted in a blinded manner to avoid bias.

### Bioinformatics analysis for gene interaction

The interaction between lncRNA and miRNA was predicted using bioinformatics tools: starBase 3.0 and miRDB. Putative targets of miRNA were identified using TargetScan and miRDB.

### Statistical analysis

To evaluate the statistical significance of molecular changes and clinical factors, we applied the Mann-Whitney U test for continuous variables and the Chi-Square test for categorical variables. The association between the molecular changes and clinical/demographic data was examined using Pearson’s correlation analysis. For the identification and optimization of potential biomarkers, we employed the Least Absolute Shrinkage and Selection Operator (LASSO) method in conjunction with logistic regression. To assess the discriminatory power of our biomarkers, Receiver Operating Characteristic (ROC) curve analysis was conducted, including the computation of the Area Under the Curve (AUC) along with 95% confidence intervals to evaluate the accuracy of the predictive models. For the analysis of animal survival data, we employed the Kaplan-Meier survival analysis complemented by the Log-rank test to compare survival curves among different groups.

## RESULTS

### Differences in plasma lncRNA profiles across ethnic groups

We quantified 43 lung cancer-associated lncRNAs in plasma from a cohort of 92 NSCLC patients and 76 cancer-free individuals using ddPCR (Supplementary Table 1) (14, 17, 18). Among the analyzed lncRNAs, ten (H19, HOTAIR, MALAT1, MEG3, MEG8, NEAT1, PVT1, RMRP, SNHG1, and TUG1) were consistently detectable in the plasma samples. Out of these, seven showed varied expression levels between lung cancer patients and control individuals. Particularly, H19, NEAT1, SNHG1, and TUG1 were significantly more expressed in lung cancer patients from both AA and WA lung cancer patients compared to those without cancer (Figure 1A, Supplementary Table 2). MALAT1 and PVT1 levels were mostly higher in AA lung cancer patients, while RMRP was notably elevated in WA lung cancer patients (All p<0.05). Increased levels of MALAT1, RMRP, NEAT1, and SNHG1 were significantly linked to smoking pack years, and elevated MALAT1, RMRP, and SNHG1 levels correlated with advanced NSCLC stages. There was no correlation among the lncRNAs (Supplementary Table 3).

**Figure 1.**
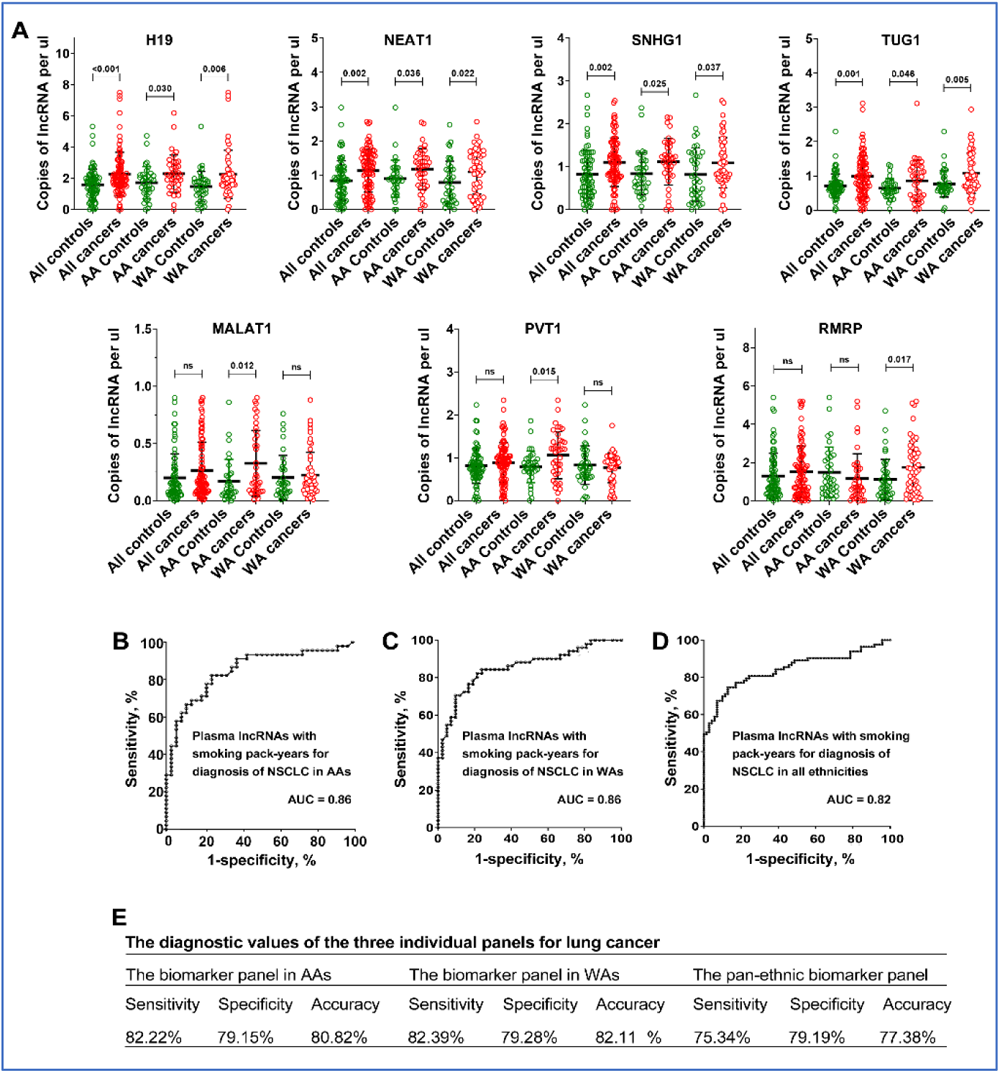
Differential expression of lncRNAs in plasma and diagnostic performance for lung cancer by ethnicity. A. Expression levels of lncRNAs in patient plasma: Scatter plots showing the plasma levels of seven lncRNAs - H19, NEAT1, SNHG1, TUG1, MALAT1, PVT1, and RMRP – in AA and WA individuals. Red dots represent control individuals and green dots represent lung cancer patients. The black lines indicate mean expression levels. Statistical significance was determined using the Mann-Whitney U test; ’ns’ denotes not significant, and p-values are shown above the plots for significant comparisons. B. Diagnostic performance for lung cancer in AAs: Receiver Operating Characteristic (ROC) curve for a biomarker panel including MALAT1, PVT1, SNHG1, and smoking pack-years, showing a high Area Under the Curve (AUC) of 0.86, indicating strong diagnostic ability for lung cancer in AA patients. C. Diagnostic performance for lung cancer in WAs: ROC curve for a biomarker panel comprising RMRP, H19, NEAT1, and smoking pack-years, with an AUC of 0.87. D. ROC curve for a pan-ethnic biomarker panel (SNHG1, NEAT1, TUG1, and smoking pack-years) applicable across all ethnicities, indicating a good diagnostic performance with an AUC of 0.82, which is lower than the ethnic-specific panels (All p<0.05). E. The diagnostic values of the three individual panels.

### The diagnostic utility of plasma lncRNAs for lung cancer in AAs

Given the differential expression of seven lncRNAs in lung cancer patients compared to cancer-free controls, we employed the LASSO (Least Absolute Shrinkage and Selection Operator) method combined with logistic regression to identify potential plasma biomarkers for lung cancer across various ethnic groups. For AAs, the most accurate prediction for lung cancer was achieved by combining three lncRNAs (MALAT1, PVT1, and SNHG1) with smoking pack-years. This combination yielded an AUC of 0.86, effectively distinguishing AA cancer patients from healthy AAs, with a sensitivity of 82%, a specificity of 79%, and an accuracy of 81% (Figure 1B). For WAs, a combination of three lncRNAs (RMRP, H19, and NEAT1) along with smoking pack-years achieved an AUC of 0.87, enabling the diagnosis of NSCLC with 83% sensitivity and 79% specificity, and 81% accuracy (all p < 0.05) (Figure 1C). In the context of pan-ethnic diagnosis, a panel comprising plasma SNHG1, NEAT1, TUG1, and smoking pack-years exhibited the strongest universal diagnostic performance, yielding an AUC of 0.82, a sensitivity of 76%, a specificity of 79%, and an accuracy of 82% (Figure 1D) (Figure 1E). Interestingly, the pan-ethnic biomarker panel displayed significantly lower sensitivity for AAs and WAs when compared to their respective individual biomarker panels (76% vs. 82% for AAs and 83% for WAs, p < 0.05), while maintaining a similar level of specificity (79%) (Figure 1E). When used as panels, the diagnostic effectiveness of the plasma lncRNA biomarkers was consistent across multiple variables, including patient age, sex, and the histological types of lung tumors, with the exception of lung cancer stages.

### Overexpression of MALAT1 is associated with monocyte chemoattractant protein-1 (MCP-1), a chemokine, in AA patients with lung cancer

lncRNAs have been proposed to play a role in the immune mechanisms involved in tumorigenesis (25). To explore whether MALAT1 contributes to immune responses in lung cancer among AAs, we evaluated cytokines in the same plasma sets and compared these with the results for lncRNAs. MCP-1 and IL-10 levels were specifically elevated in AA lung cancer patients (Figure 2A and Supplementary Table 4). In contrast, WA patients exhibited increased levels of IL-6. Notably, a significant correlation was found between MALAT1 and MCP-1 levels in the plasma of lung cancer patients from the AA population (r=0.935, p=0.001) (Figure 2B), a correlation not observed in the WA population (r=0.229, p=0.096) (Figure 2 B and C). A high expression of PVT1 is also observed in lung cancer among AA patients; however, it is not associated with the cytokines, including MCP-1 (r=0.201, p=0.221) (Figure 2D).

**Figure 2.**
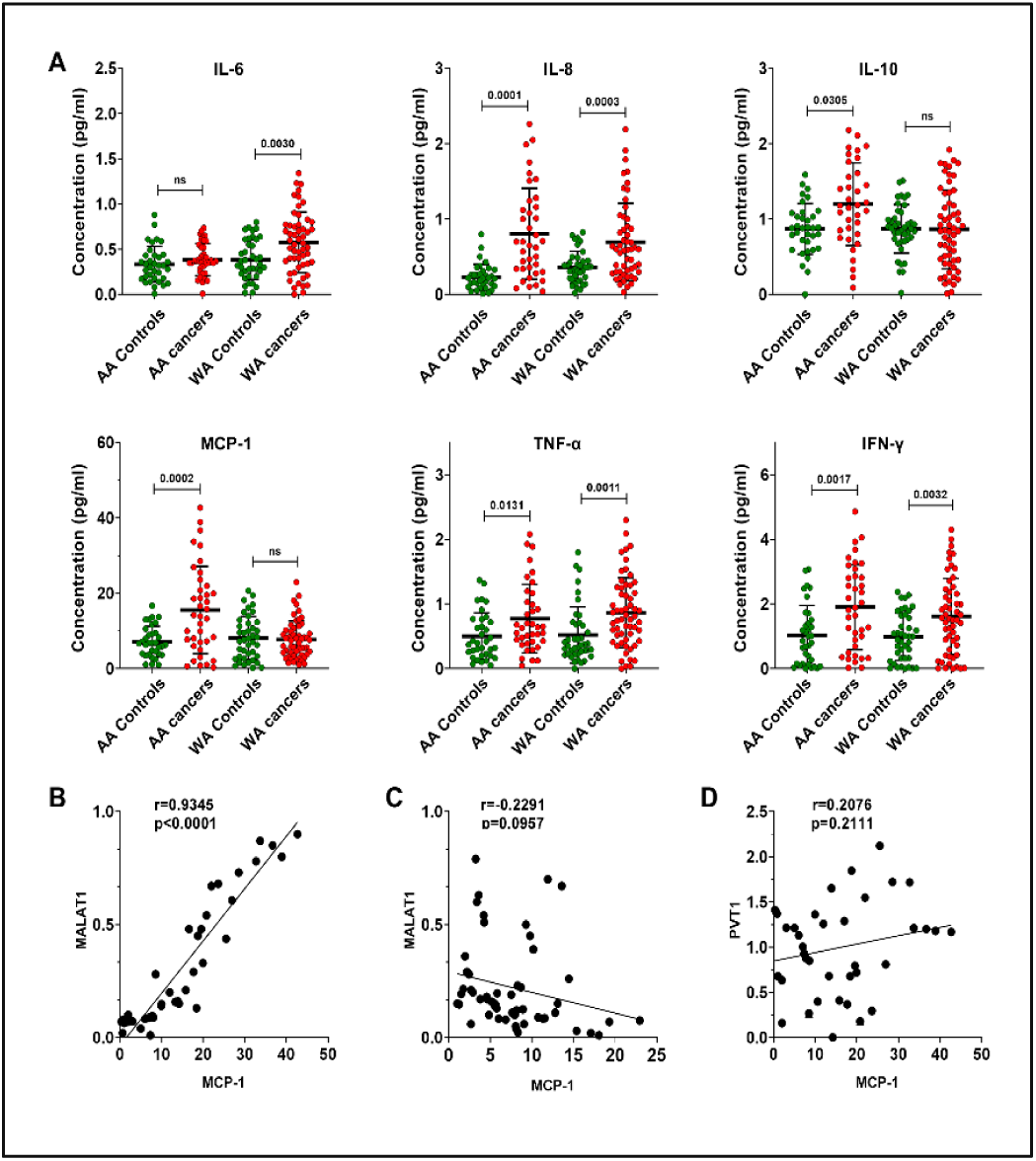
Comparative analysis of cytokine levels in plasma of AA and WA lung cancer patients. A. Elevated IL-6 levels were observed in WA lung cancer patients compared to their controls. Both AA and WA lung cancer groups showed increased expression of IL-8, IFN-γ, and TNF-α relative to their respective control groups. Notably, IL-10 and MCP-1 were exclusively upregulated in the AA lung cancer cohort. B. A strong positive correlation (Pearson’s R=0.9345, P < 0.0001) was found between plasma MALAT1 and MCP-1 levels in AA lung cancer patients. Each dot on the scatter plot corresponds to an individual patient’s paired biomarker expression, with MALAT1 expression on the y-axis and MCP-1 concentration on the x-axis. C. No correlation existed between plasma MALAT1 and MCP-1 levels in WA lung cancer patients. D. No correlation existed between plasma PVT1 and MCP-1 levels in AA lung cancer patients.

### MALAT1 and MCP-1 are both highly expressed in lung tumor tissues and cancer cell lines from AA patients

Since upregulation of MALAT1 is associated with elevated level of MCP-1 in plasma of AA lung cancer patients. We broadened our study to analyze MALAT1 and MCP-1 in lung tumor tissues and adjacent non-cancerous lung tissues of 46 NSCLC patients, including 23 AAs and 23 WAs. Both MALAT1 and MCP-1 levels were elevated in lung tumors compared to normal controls (Figure 3A and Supplementary Table 5). While MALAT1 and MCP-1 levels were higher in lung tumor specimens from WA lung cancer patients compared to their normal tissue counterparts (p=0.043 and 0.041, respectively), a more pronounced increase was observed in lung tumors of AA patients relative to those in WAs (p=0.001 and 0.001, respectively) (Figure 3A-B). Furthermore, a positive correlation between MALAT1 and MCP-1 levels was also noted in the tissues (r=0.801, p<0.001) (Figure 3C).

**Figure 3.**
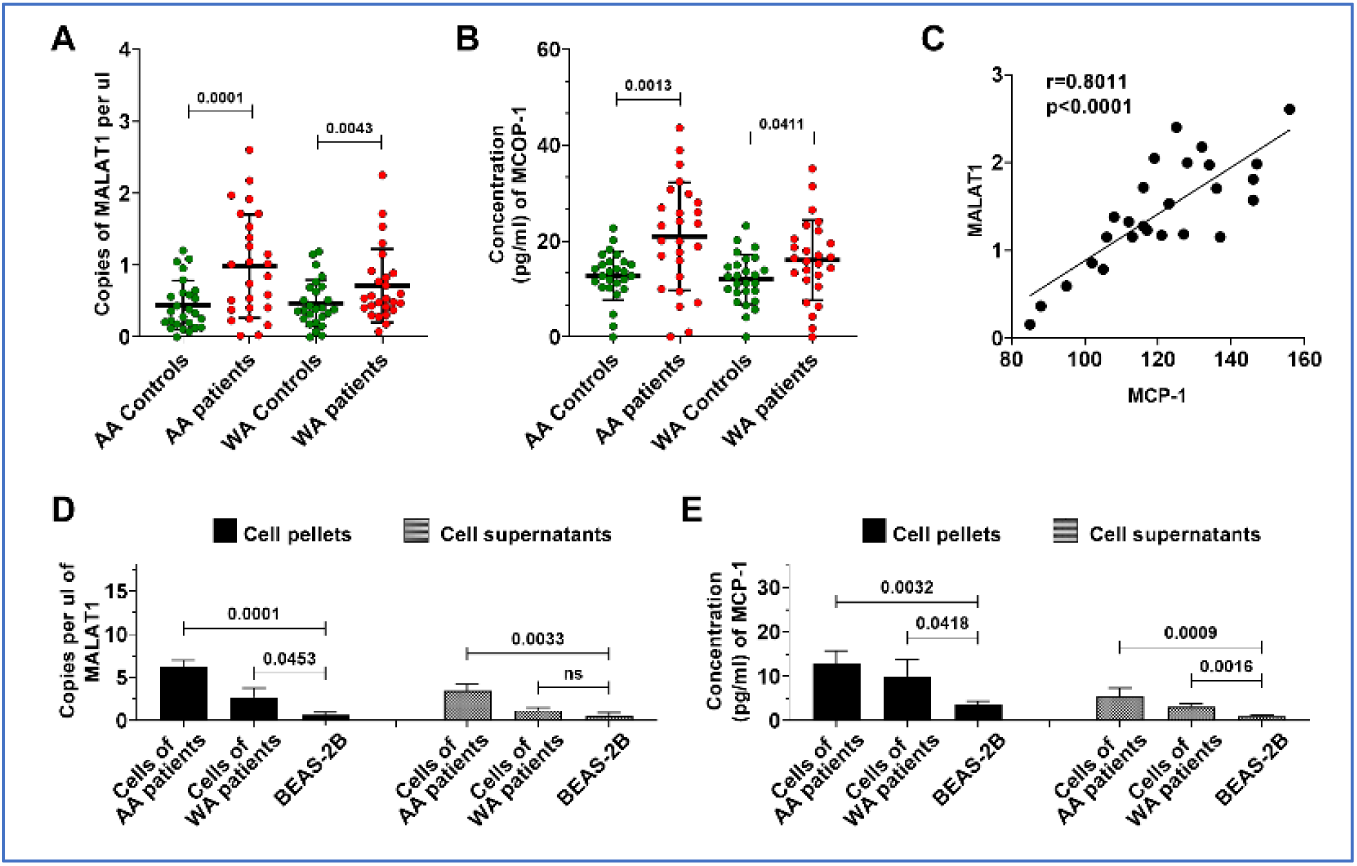
Differential expression and correlation of MALAT1 and MCP-1 in lung tumor tissue and cancer cell lines by racial demographics. A and B. Significantly higher levels of MALAT1 and MCP-1 in lung tumor specimens compared to normal tissue controls. Furthermore, a notable increase in both MALAT1 and MCP-1 is observed in AA patients as opposed to WA patients, respectively (All p<0.05). C. A strong positive correlation between MALAT1 and MCP-1 levels in lung tumor tissues of AA patients, with a Pearson’s correlation coefficient (r=0.8011, p<0.0001). D. Bar graphs showing significantly increased levels of MALAT1 in cellular pellet and supernatant samples of AA patients compared to those from WA patients and normal cells. E. Elevated concentrations of MCP-1 in cellular pellet and supernatant samples of AA patients compared to those from WA patients and normal cells.

We further corroborated these results using six NSCLC lung cancer cells and their supernatants. Echoing our observations in lung tumor tissues, both MALAT1 and MCP-1 were found to be elevated in cancer cell lines and their supernatants, relative to normal bronchial epithelial cells (Figure 3D-E, Supplementary Figure 1). Furthermore, five lung cancer cell lines including A549, H513, H1373, and H1385, and H520 exhibited higher levels of MALAT1 and MCP-1 compared to H460 and Beas-2B cells (Supplementary Figure 1). Interestingly, the H513, H1373, and H1385 lung cancer cell lines, originating from an AA patient, exhibited a higher level of MALAT1 and MCP-1 levels in both cells and their supernatants compared to other three cell lines (A549, H460, and H520) developed from WA lung cancer patients (Figure 3D-E). Considering that supernatants may resemble plasma in vivo, this finding reflects our above observations in plasma samples from AA lung cancer patients, who also demonstrated increased MALAT1 levels in AA patients diagnosed with lung cancer.

### Upregulation of MALAT1 enhances tumor-associated macrophage presence and alters the dynamics of the tumor immune microenvironment (TIME)

MCP-1 is suggested to influence tumor growth and progression by attracting and activating tumor-associated macrophages within the TIME (26). Given the positive correlation between MALAT1 and MCP-1 levels in tissues, we use immunohistochemistry (IHC) to evaluate macrophage markers (CD68, CD163, CD206, and iNOS), in addition to MCP-1 and KI-67, in the same tissue samples, to correlate these findings with changes in MALAT1. Markers for M2 macrophages, including CD68, CD163, and CD206, were found to be higher in lung tumor tissues compared to adjacent non-tumor lung tissues, regardless of histological types, including AC and SCC (Fig 4A-B). Additionally, levels of MCP-1 and Ki-67 were also elevated in tumor tissues(Fig 4A-B). Conversely, the M1 macrophage marker, iNOS, showed no significant difference between tumor and non-tumor lung tissues (Fig 4A-B). A positive link was identified between MALAT1 and MCP-1 levels and the presence of CD68, CD163, and CD206 in AA patient tissues (Figure 4C-H). In the tumor tissues of AA patients, a significant correlation was found between CD206 and CD163, markers indicative of M2 macrophages (Figure 4I), as opposed to iNOS, a marker for M1 macrophages. These IHC findings suggest that increased MALAT1 expression in AA lung cancer patients may trigger MCP-1 activation, thereby enhancing tumor-associated macrophage presence and altering the tumor microenvironment dynamics.

**Figure 4.**
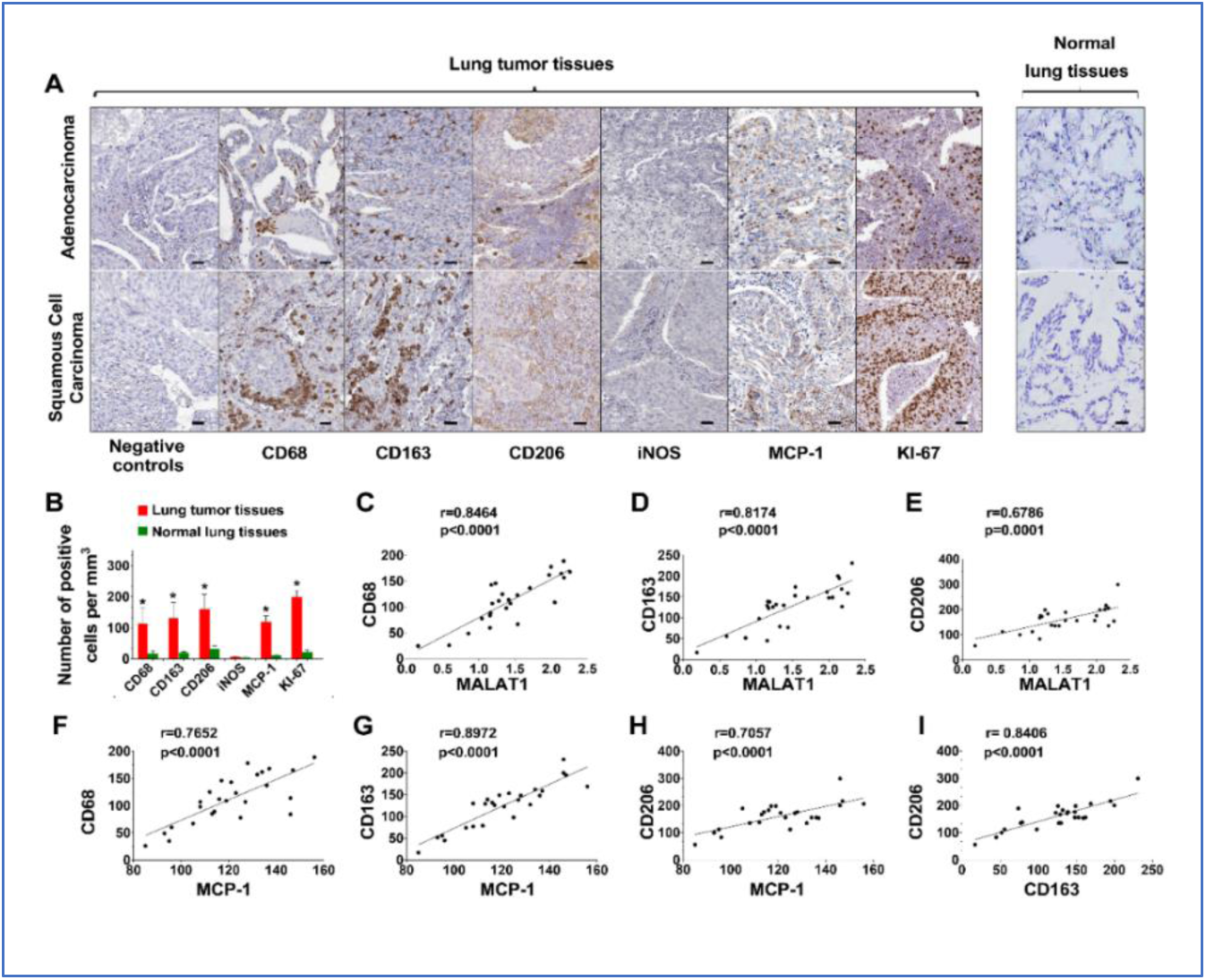
IHC analyses comparing lung tumor tissues to normal lung tissues for CD68, CD163, CD206, iNOS, MCP-1, and Ki-67 with negative controls ensuring specificity. A. In tumor tissues, including AC and SCC, an increased expression of CD68, CD163, CD206, MCP-1, and Ki-67 is observed, whereas iNOS levels remain unchanged. Bar = 50 μm. B. Quantitative analysis and correlation of the results presented in A, indicating the elevated expression of CD68, CD163, CD206, MCP-1, and Ki-67 within tumor tissues. *, all <0.001. C-H. Significant positive correlations between MALAT1 expression and M2 markers, and between MCP-1 and M2 markers, specifically in samples from AA lung cancer patients (All p<0.001). I. A significant positive correlation between the levels of CD206 and CD163, which are markers for M2 macrophages, in tissue samples from AA lung cancer patients (p<0.001).

### Upregulation of MALAT1 promotes the tumorigenicity of lung cancer by activating MCP-1 in NSCLC cells

We further functionally evaluated whether increasing MALAT1 expression could enhance the *in vitro* tumorigenic capabilities of cancer cells. Forced MALAT1 expression led to increased cell proliferation, invasion, and migration in the H513 and H460 cells (Figures 5A-C). By contrast, reducing MALAT1 levels in the cells markedly diminished the malignant behavior of NSCLC cells (Supplementary Figure 2). Furthermore, an increase in MALAT1 expression was correlated with a marked elevation in MCP-1 levels in A549, H460, and H520c ells, which generally display lower MCP-1 expression (Figures 5D). In contrast, diminishing MALAT1 expression in H513, H1373, and H1385 cells, characterized by relatively higher MCP-1 expression, resulted in a substantial reduction in MCP-1 levels (Figures 5E). The findings suggest that MALAT1 may regulate MCP-1 expression and enhance malignant behaviors, while its knockdown significantly reduces these characteristics in lung cancer cells.

**Figure 5.**
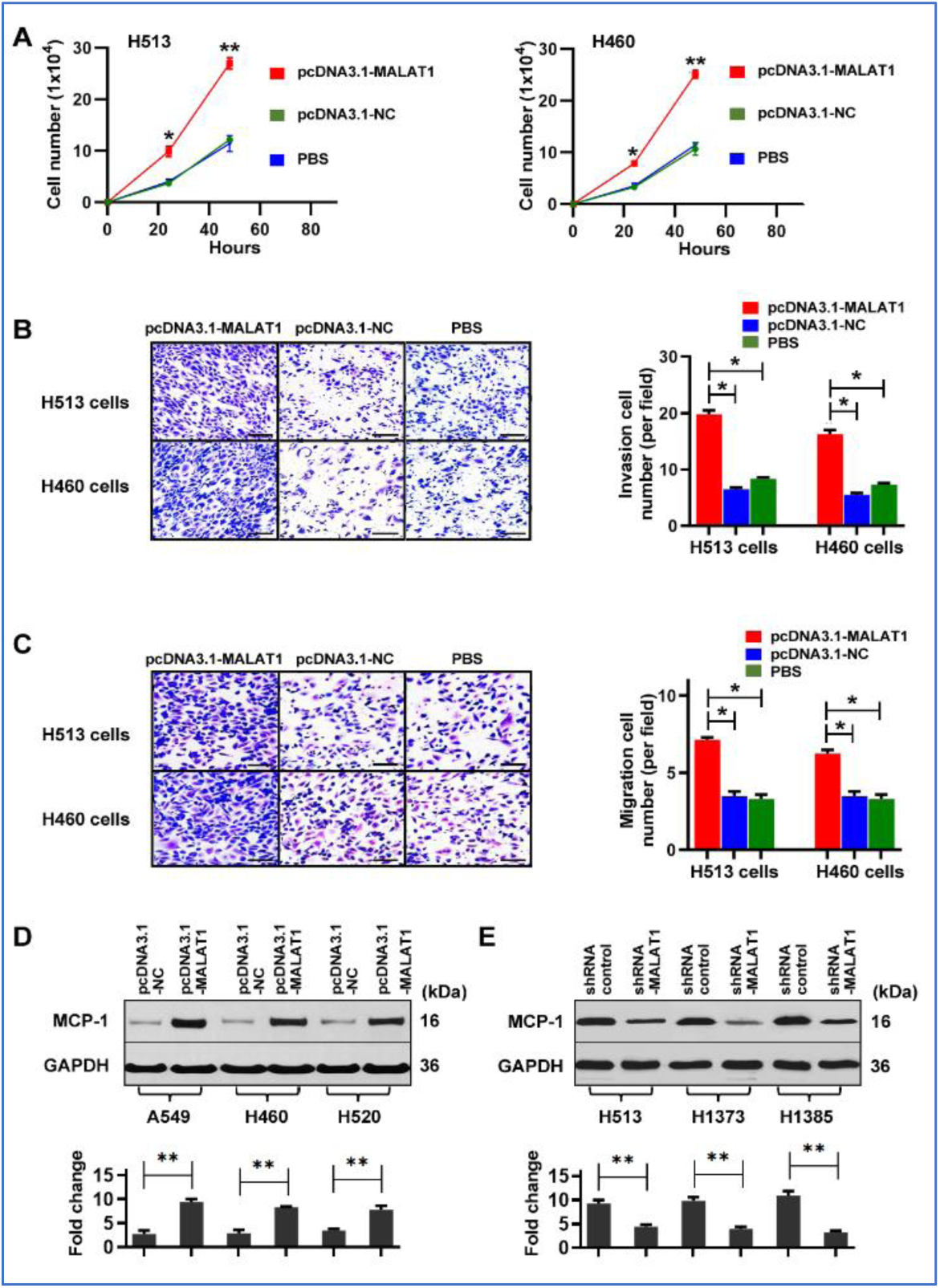
Effects of MALAT1 modulation on NSCLC cell behavior and MCP-1 levels. A. Treatment of H513 and H460 cancer cells with pcDNA3.1-MALAT1 for MALAT1 overexpression, compared to pcDNA3.1-NC negative control and PBS control, led to significantly increased cell proliferation. *, all p<0.05; **, all p<0.01. Error bars represent the standard deviation of the mean from three experiments. B. Representative images and quantitative analysis of invasive H513 and H460 cells through a Matrigel-coated membrane. MALAT1 overexpression (pcDNA3.1-MALAT1) substantially increased the invasive capabilities of the cells relative to the negative (pcDNA3.1-NC) and PBS controls, as shown on the right bar chart. Bar = 50 μm. *, all p<0.05. C. Microscopic images and quantitative analysis of the migration of H513 and H460 cells. Overexpression of MALAT1 (pcDNA3.1-MALAT1) led to a marked increase in cell migration compared to negative (pcDNA3.1-NC) and PBS controls, demonstrated by the bar graph on the right. Bar = 50 μm. *, all p<0.05. D. Western blot analysis reveals that increased MALAT1 expression is associated with higher MCP-1 protein levels in A549, H460, and H592 NSCLC cells, known for their typically lower endogenous MCP-1 expression. Below, the mean (±SD) densitometric analysis of the western blots is presented, with fold changes normalized against the housekeeping gene GAPDH. **p<0.01. E. Western blot analysis demonstrates that reducing MALAT1 expression via siRNA treatment in H513, H1373, and H1385 cells, which usually have higher MCP-1 expression, results in a notable decrease in MCP-1 protein levels. **p<0.01.

### miR-206 acts as a direct downstream target of MALAT1

Functioning as ’sponges’ or competitive endogenous RNAs (ceRNAs), lncRNAs can modulate gene expression by influencing the stability and translation process (25). To investigate the regulatory mechanisms of MCP-1 by MALAT1, we employed Starbase 2.0 and miRDB to identify miRNAs that potentially bind to MALAT1 and target MCP-1. Bioinformatics analysis suggested that MALAT1 could directly targets miR-206, which in turn influences MCP-1 expression (Figure 6A-B).

**Figure 6.**
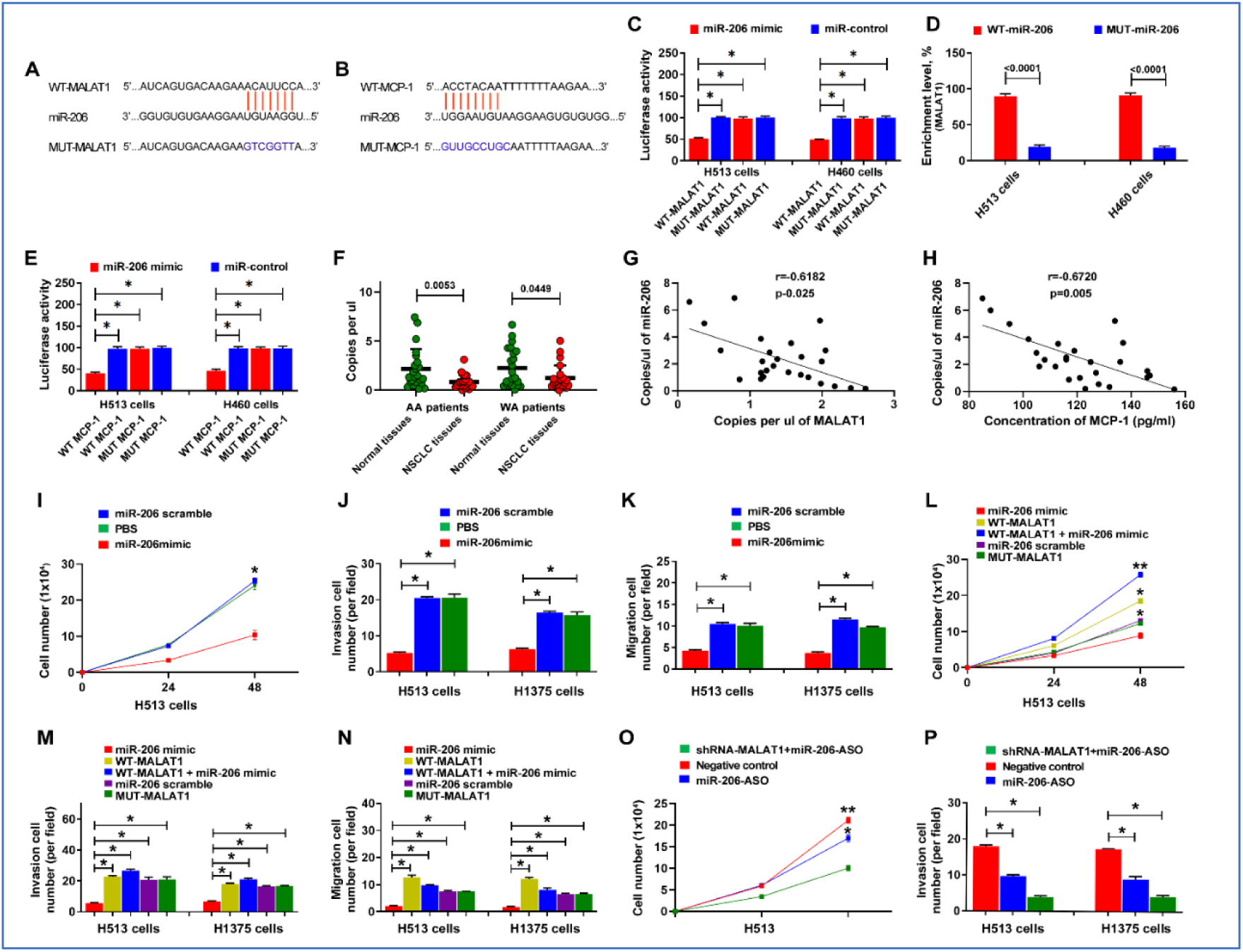
MALAT1 directly targets miR-206, influencing MCP-1 expression, and contributes to the lung tumorigenesis. A-B. Bioinformatics analysis suggested that MALAT1 may directly target miR-206, and in turn, miR-206 may directly target MCP-1. C. Dual luciferase reporter assay results showing the relative luciferase activity in H513 and H460 lung cancer cells. Cells were co-transfected with either WT-or MUT-MALAT1 constructs along with miR-206 mimic or control. Blue bars represent WT-MALAT1 constructs; red bars represent MUT MALAT1 constructs. A reduction in luciferase activity is observed with WT-MALAT1 and miR-206 mimic co-transfection. *, all p<0.05. D. Enrichment of miR-206 in the RNA pull-down assay with biotinylated MALAT1 constructs. Blue bars represent the enrichment after pull-down with WT-MALAT1 constructs; red bars represent enrichment with MUT-MALAT1 constructs. E. Dual luciferase reporter assay results showing the effect of miR-206 on the 3′UTR of MCP-1. Constructs with WT or MUT 3′UTR of MCP-1 were co-transfected into lung cancer cells with miR-206. Decreased luciferase activity with WT 3′UTR and miR-206 mimic suggests that miR-206 binds to MCP-1 3′UTR, leading to downregulation of the luciferase reporter. *, all p<0.05. F. The levels of miR-206 in lung cancer tissues compared to non-cancerous lung tissues. Lower levels of miR-206 expression were observed in lung cancer tissues. G-H. Scatter plot depicting the inverse correlation between miR-206 copy number and expression levels of MALAT1 and MCP-1 in tumor tissues. Each dot represents a tissue sample, with the X-axis showing MALAT1 or MCP-1 levels and the Y-axis showing miR-206 copy number. I. Proliferation assay showing cell numbers of lung cancer cells treated with either miR-206 mimic or miR-206 scramble control over a 48-hour period. *, p<0.01. J. Invasion assay results for H513 and H1385 cells treated with PBS, miR-206 mimic, or miR-206 scramble, indicating reduced invasive capacity upon miR-206 mimic treatment. *, all p<0.05. K. Migration assay results for H513 and H1385 upon miR-206 mimic treatment. *, all p<0.05. L. Cell proliferation assays for H513 and H1385 cells treated with miR-206 mimic, WT-MALAT1, WT-MALAT1 plus miR-206 mimic, miR-206 scramble, or MUT-MALAT1 over 48 hours. *, all p<0.05; **, all p<0.01. M. Invasion assay for cancer cells treated with the above conditions, showing the mitigating effect of WT-MALAT1 overexpression on miR-206-induced suppression of invasion. *, all p<0.05. N. Migration assay for cancer cells treated with the above conditions, showing the mitigating effect of WT-MALAT1 overexpression on miR-206-induced suppression of migration. *, all p<0.05.. *, all p<0.05. O. Proliferation assay of H513 cells treated with sh-MALAT1 plus miR-206 ASO, negative control, or miR-206-ASO alone, showing restored proliferation with miR-206 ASO treatment. *, all p<0.05; **, all p<0.01. P. Invasion assay for H513 and H1385 cells under the same treatment conditions as in (O), demonstrating the reversion of sh-MALAT1 tumor-suppressive effects by miR-206 ASO. *, all p<0.05. Error bars represent the standard deviation of the mean from three experiments.

We used the Dual Luciferase Reporter Assay to validate the bioinformatics analysis. Vectors harboring either the WT-or MUT-MALAT1 were co-transfected into H153 cancer cells along with the miR-206 mimic and control. The co-transfection resulted in a significant reduction in luciferase activity when the WT MALAT1 was paired with miR-206, thereby confirming their interaction (Figure 6C). Further validation was performed by using an RNA pull-down assay with biotin-labeled constructs. A significant enrichment of miR-206 in complexes with MALAT1 was observed (Figure 6D). Furthermore, the 3′UTRs of MCP-1 with predicted miR-206-binding sites and their mutant versions were co-transfected into lung cancer cells with miR-206 mimic and control. The cells transfected with the WT MCP-1 3′UTR and miR-206 mimic showed decreased luciferase activity, suggesting miR-206 could binds to the 3′UTR of MCP-1, leading to the degradation or translational repression of the luciferase reporter mRNA (Figure 6E). We also assessed the levels of miR-206 in lung cancer tissue samples, which had been previously analyzed for MALAT1 and MCP-1 expression. miR-206 was expressed at lower levels in lung cancer tissues compared to their corresponding non-cancerous lung tissues (Figure 6F). Additionally, an inverse relationship was observed between the copy number of miR-206 and the expression levels of both MALAT1 and MCP-1 (r=-0.618 and -0.672, respectively, all p<0.05) (Figure 6G-H). Altogether, MALAT1 might directly interact with miR-206, thereby regulating MCP-1 and playing a role in the development and progression of lung tumorigenesis.

### MALAT1 enhances malignant behaviors of lung cancer cells by targeting miR-206

We further investigate MALAT1’s role in lung carcinogenesis through its intricate regulatory interaction with miR-206. Overexpression of miR-206 markedly impeded proliferation, migration, and invasion of the cancer cells (Figures 6I-K) (Supplementary Figure 3). Intriguingly, simultaneous transfection of cells with MALAT1 and miR-206 mimics counteracted the proliferation, migration, and invasion caused by miR-206 mimics (Figures 6L-N) (Supplementary Figure 3). We performed rescue experiments to elucidate the role of the miR-206/MCP-1 axis in MALAT1 downregulation-driven attenuation of oncogenic traits. Post miR-206 ASO administration, the sh-MALAT1-induced reductions in cell proliferation, migration, and invasion were substantially reversed (Fig 6 O-P) (Supplementary Figure 3). Taken together, these results compellingly suggest that MALAT1 enhances the malignant behaviors of NSCLC cells by targeting and downregulating miR-206.

### MALAT1 promotes growth of lung tumor in xenograft animal models

The mice were randomly divided into two groups: Group 1 received subcutaneous injections of H460-Luc cells transfected with the empty pcDNA3.1(+) vector (NC), while Group 2 was injected with H460-Luc cells transfected with pcDNA3.1-MALAT1. By the end of the third week, tumors derived from MALAT1-expressing cells in group 2 were notably larger, with an average volume of 68.67±31.02 mm ³, in contrast to the tumors in group 1, which averaged 26.11±15.86mm³ (p=0.015) (Figure 7A-B). These tumors also exhibited higher levels of MCP-1, CD68, CD163, and KI-67 compared to tumors from the control group (Figure 7C). These findings in the xenograft mouse model corroborate our in vitro observations, further confirming the driver role of MALAT1 in lung tumorigenesis.

**Figure 7.**
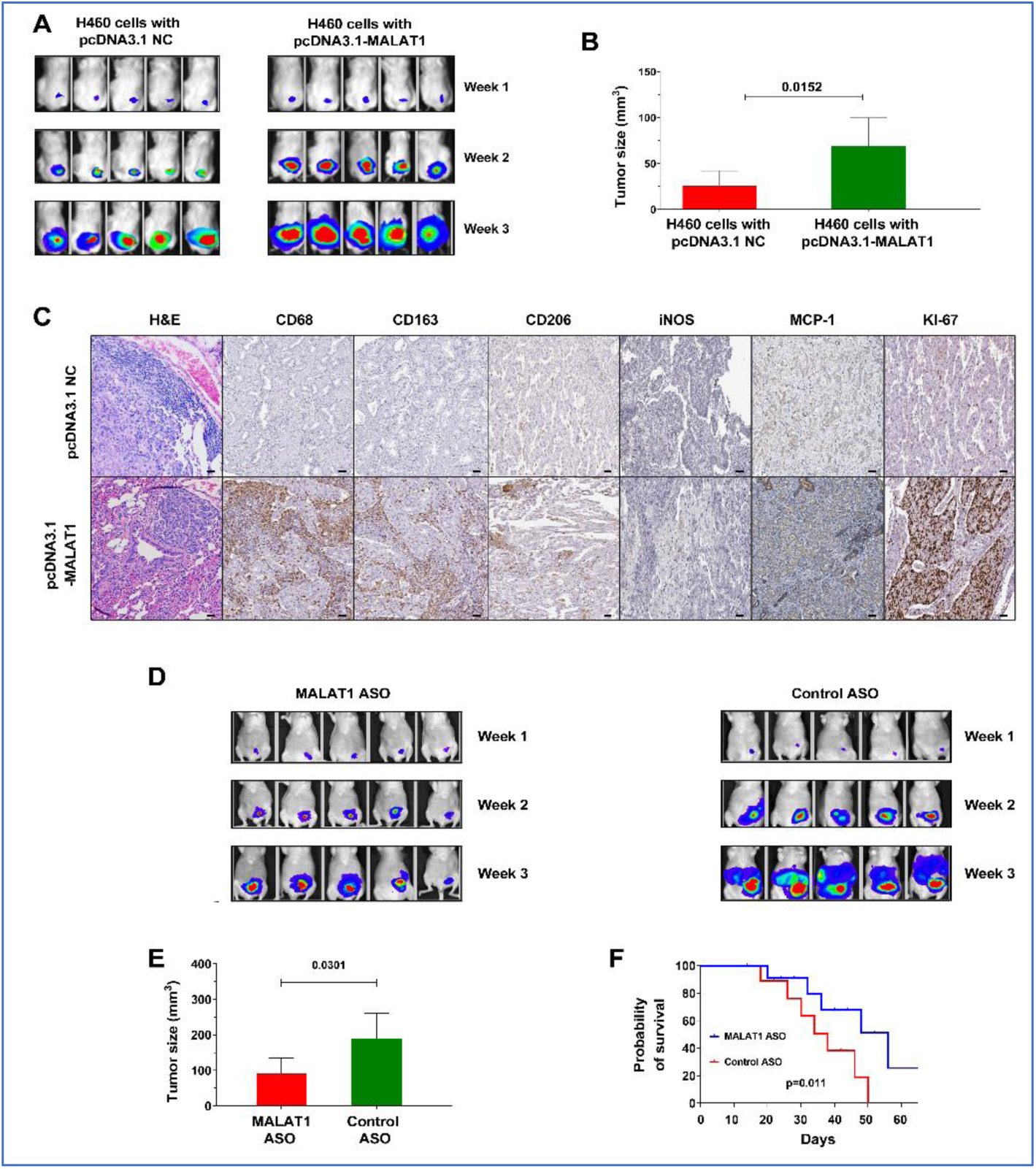
Impact of MALAT1 on lung cancer development and progression and the therapeutic efficacy of MALAT1 ASO in xenograft mouse models. A. Imaging of two groups of mice over three weeks showing the development and progression of xenograft tumors. One group received injections of H460 lung cancer cells with MALAT1 overexpression, while the other group was injected with cells containing an empty vector (pcDNA3.1 NC). An early onset of tumor development is observed in the group with H460 lung cancer cells with MALAT1 overexpression, indicating a MALAT1-driven increase in tumorigenicity. B. Representative images of tumors excised from both groups of mice at 28 days post-cell injection. Tumors from the group received injections of H460 lung cancer cells with MALAT1 overexpression are visibly larger, corroborating the bioluminescence data. C. Hematoxylin and eosin (H&E) staining displays the histological structure of lung tumors. Immunohistochemical staining show that MALAT1-overexpressing tumors exhibit enhanced levels of MCP-1, CD68, CD163, and KI-67 when compared to control. Bar = 50 μm. D. Bioluminescence imaging of mice post-treatment showing tumor regression in the group treated with MALAT1 ASOs compared to the control ASO group. E. Measurement of tumors at the end of the treatment period, highlighting a significant decrease in tumor volume in the MALAT1 ASO-treated group versus control (p=0.031). F. Survival curves for the two groups of mice, one treated with MALAT1 ASO and the other with control ASO. The curve demonstrates a significant survival advantage in the MALAT1 ASO-treated group (Kaplan-Meier survival analysis).

### Effective antitumor effects of MALAT1-targeting ASOs in vivo

To explore the impact of MALAT1 inhibition on tumor growth in vivo, A549-Luc cells, known for high MALAT1 expression, were subcutaneously injected into mice. Within 10 days, tumors developed in all injected mice. These mice were then treated three times per week with MALAT1 ASO or control ASO, by injecting directly into the tumors. Tumors treated with MALAT1 ASO exhibited a significantly larger reduction in volume compared to the control group (189.58±70.59 mm³ vs. 90.75±44.47 mm³, p=0.031) (Figure 6 D-E). Furthermore, mice receiving MALAT1 ASO exhibited a longer survival period than those treated with control ASO (p<0.05) (Figure 6F). This outcome implies that MALAT1 inhibition could effectively reduce tumor growth in vivo, highlighting its potential as a therapeutic strategy.

## DISCUSSION

AA populations experience higher incidence and mortality rates from lung cancer compared to other ethnic groups. Exploring the molecular and biological mechanisms behind these disparities is crucial for developing effective diagnostic and therapeutic strategies aimed at addressing the inequalities in lung cancer outcomes. Our comprehensive study demonstrated that in AA populations, the upregulation of MALAT1 plays a crucial role in the development and progression of lung cancer. By modulating miR-206 and MCP-1, MALAT1 contributes to increased cell proliferation and cancer progression of lung cancer in AA patients. We also show the possibility that inhibiting MALAT1 may significantly diminish tumor size in animal models, thereby presenting novel diagnostic and treatment strategies for lung cancer in AAs and potentially mitigating racial disparities in lung cancer prognosis.

We found that MLATA1 level is higher in both lung tumor tissues and plasma samples of AA lung cancer patients. Consistently, lung cancer cell lines, originating from AA patients, exhibited a high level of MALAT1 in both cells and their supernatants compared to other three cell lines developed from WA lung cancer patients. Considering that supernatants may resemble plasma in vivo, this finding reflects our above observations in plasma samples from AA lung cancer. Notably, the incorporation of lncRNA profiles, along with smoking history, resulted in a high diagnostic accuracy (89%) in AA patients. Our findings suggest that lncRNA-based liquid biopsy could be a significant step forward in the early detection and intervention of lung cancer in racially diverse populations.

MALAT1’s roles are diverse and complex, influencing a range of cellular processes fundamental to tumor development, growth, invasion, and metastatic spread (24, 27–30). Notably, while numerous studies have documented the oncogenic function of MALAT1 in various tumors, its specific role in lung cancer among AAs had not been previously investigated. It is well accepted that a dynamic and reciprocal relationship develops between cancer cells and components of the TIME that supports cancer cell survival, local invasion, and metastatic dissemination(26). Furthermore, upregulation of MCP-1, a potent monocyte-attracting chemokine, in the TIME could drive cancer development and progression via recruiting M2 macrophages within tumor microenvironment (26). Here we observe a rise in MALAT1 expression, correlating with increased levels of MCP-1 and CD68, CD163, CD206, indicative of M2 macrophages in tumor tissues of AA lung cancer patents. Forced MALAT1 expression in cancer cells and xenograft models led increased cell growth, invasiveness, and movement, along with higher levels of MCP-1, CD68, CD163, CD206, and KI67. The MALAT1-reguated TIME can facilitate the breakdown of cell-cell adhesion, and enhances migratory properties, all of which collectively contribute to tumor growth and metastatic dissemination. Therefore, MALAT1 affects MCP-1 expression and macrophage activity, and enhance TIME and tumorigenesis. Furthermore, our use of antisense oligonucleotides to inhibit MALAT1 expression led to a significant reduction in tumor sizes in animal models, suggesting the therapeutic potential of targeting lncRNAs. Previous study suggest that MALAT1 overexpression can drives tumor progression with a lesser impact on tumor growth in lung adenocarcinoma (31). Our current study, employing xenograft mouse models, cell lines, and human plasma and tissue samples, found that MALAT1 upregulation affects both tumor growth and progression in lung cancer within the AA population. Significantly, our study further clearly demonstrated that, mechanistically, MALAT1 interacts with miR-206 to modulate MCP-1 levels, thereby influencing the TIME and promoting tumorigenesis in AAs.

While our study sheds light on important aspects of lncRNAs in lung cancer pathogenesis among AAs. However, a large population to prospectively validate the diagnostic performance in lung cancer is needed. Furthermore, our *in vitro* and *in vivo* studies need to be substantiated through clinical trials to determine the safety and efficacy of targeting MALAT1 in human subjects with a specific focus on AA populations. In addition, exploring the roles of MALAT1’s contribution to treatment resistance is also imperative.

In conclusion, our study highlights the significant role of lncRNAs, particularly MALAT1, in the racial disparities observed in lung cancer. By unraveling the complex mechanisms involving MALAT1, miR-206, and MCP-1, we provide a new perspective on the molecular basis of these disparities. Our results could potentially lead to the development of innovative diagnostic and therapeutic approaches, thereby influencing the management and outcomes of lung cancer across diverse racial groups.

## Data Availability

All data produced in the present work are contained in the manuscript

https://www.medschool.umaryland.edu/profiles/jiang-feng/

## Acknowledgments

We extend our sincere gratitude to the Biostatistics Shared Service at the University of Maryland Marlene and Stewart Greenebaum Cancer Center for their invaluable contribution in conducting the statistical analysis for this study.

## Author contributions

Conceptualization: FJ. Methodology: NL, PD, VH, AS, FJ. Investigation: LG, PD, FJ. Visualization: NL, PD, FJ. Funding acquisition: FJ. Project administration: FJ. Supervision: FJ. Writing – original draft: NL, PD, FJ. Writing – review & editing: NL, PD, FJ.

## Supplementary Materials

**Table S1.**
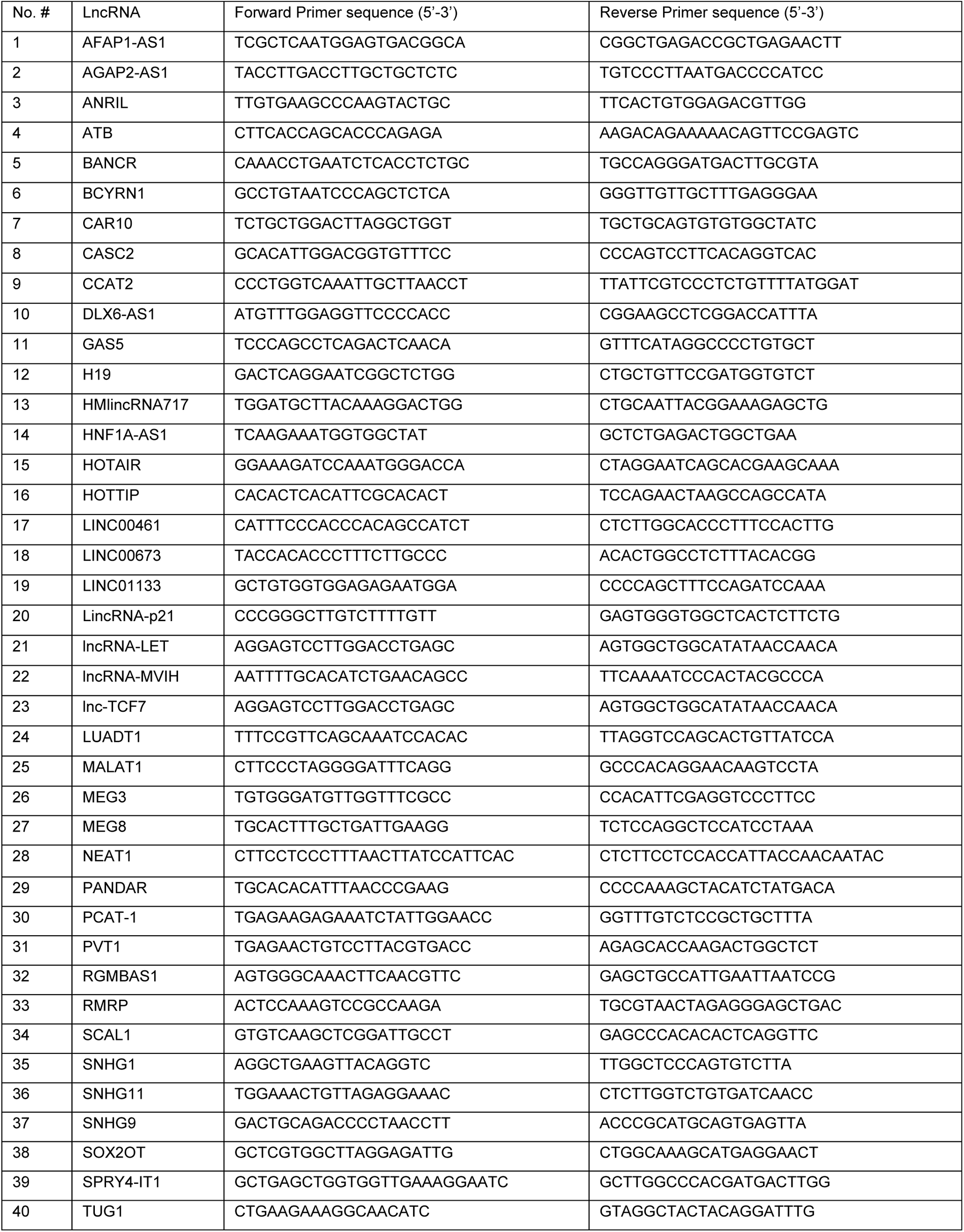
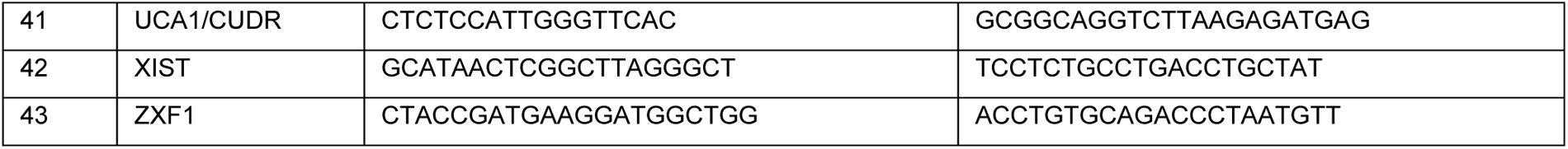
Forty-three lung cancer-associated lncRNAs were tested in this study.

**Table S2.** Mean expression levels of seven lncRNAs in lung cancer patients compared to their cancer-free counterparts.

|  | All controls | All lung cancer patients | P | AA Controls (35) | AA cancer patients | P | WA Controls | WA cancer patients | P |
| --- | --- | --- | --- | --- | --- | --- | --- | --- | --- |
| H19 | 1.601 | 2.281 | 0.001 | 1.730 | 2.302 | 0.035 | 1.481 | 2.266 | 0.006 |
| NEAT1 | 0.842 | 1.135 | 0.002 | 0.902 | 1.182 | 0.037 | 0.789 | 1.101 | 0.022 |
| SNHG1 | 0.827 | 1.103 | 0.002 | 0.837 | 1.118 | 0.023 | 0.817 | 1.093 | 0.031 |
| TUG1 | 0.710 | 0.987 | 0.001 | 0.640 | 0.862 | 0.046 | 0.773 | 1.079 | 0.005 |
| MALAT1 | 0.202 | 0.271 | 0.062 | 0.171 | 0.338 | 0.007* | 0.208 | 0.222 | 0.737 |
| PVT1 | 0.811 | 0.891 | 0.248 | 0.789 | 1.059 | 0.015* | 0.830 | 0.767 | 0.451 |
| RMRP | 1.301 | 1.523 | 0.2643 | 1.500 | 1.179 | 0.284 | 1.122 | 1.755 | 0.017 |

\*, p<0.05. The Mann-Whitney U test was used to compare the cytokines between these groups, with an alpha level of 0.05 set as the threshold for determining statistical significance.

**Table S3.** Associations between the ncRNAs and clinical and demographic data, analyzed using Pearson’s correlation coefficients.

|  | MALAT1 | RMRP | H19 | NEAT1 | PVT1 | SNHG1 | TUG1 | Age | Sex | Pack-smoking years | Types of cancer | Stage | Race |
| --- | --- | --- | --- | --- | --- | --- | --- | --- | --- | --- | --- | --- | --- |
| MALAT1 |  | 0.653 | 0.496 | 0.991 | 0.065 | -0.050 | 0.082 | 0.139 | -0.032 | 0.019 | -0.165 | 0.015 | 0.015 |
| RMRP |  |  | 0.172 | 0.858 | 0.900 | 0.058 | 0.062 | 0.052 | 0.127 | 0.027 | -0.162 | 0.013 | 0.013 |
| H19 |  |  |  | 0.362 | 0.787 | -0.039 | 0.152 | 0.060 | -0.042 | 0.200 | 0.179 | 0.173 | -0.157 |
| NEAT1 |  |  |  |  | 0.467 | 0.159 | 0.209 | 0.005 | -0.084 | 0.021 | 0.094 | -0.011 | -0.005 |
| PVT1 |  |  |  |  |  | 0.522 | 0.632 | -0.085 | -0.218 | 0.141 | -0.148 | -0.077 | 0.002 |
| SNHG1 |  |  |  |  |  |  | 0.105 | 0.105 | -0.131 | 0.016 | 0.140 | 0.026 | 0.137 |
| TUG1 |  |  |  |  |  |  |  | 0.074 | 0.086 | 0.136 | 0.233 | -0.150 | 0.066 |

**Table S4.** Cytokine levels in plasma of AA and WA lung cancer patients and their control groups.

|  | AA Controls | AA cancer patients | P | WA Controls | WA cancer patients | P |
| --- | --- | --- | --- | --- | --- | --- |
| IL-6 | 0.3334 | 0.3854 | 0.2427 | 0.3862 | 0.5755 | 0.003 |
| IL-8 | 0.2312 | 0.8039 | <0.0001 | 0.3617 | 0.6968 | 0.0003 |
| IL-10 | 0.8694 | 1.198 | 0.0305 | 0.8703 | 0.8635 | 0.9415 |
| MCP-1 | 7.18 | 15.55 | 0.0002 | 8.109 | 7.679 | 0.6962 |
| TNF-α | 0.5014 | 0.774 | 0.0131 | 0.5201 | 0.8663 | 0.001 |
| IFN-γ | 1.028 | 1.901 | 0.0017 | 0.9837 | 1.618 | 0.0032 |

**Fig. S1.**
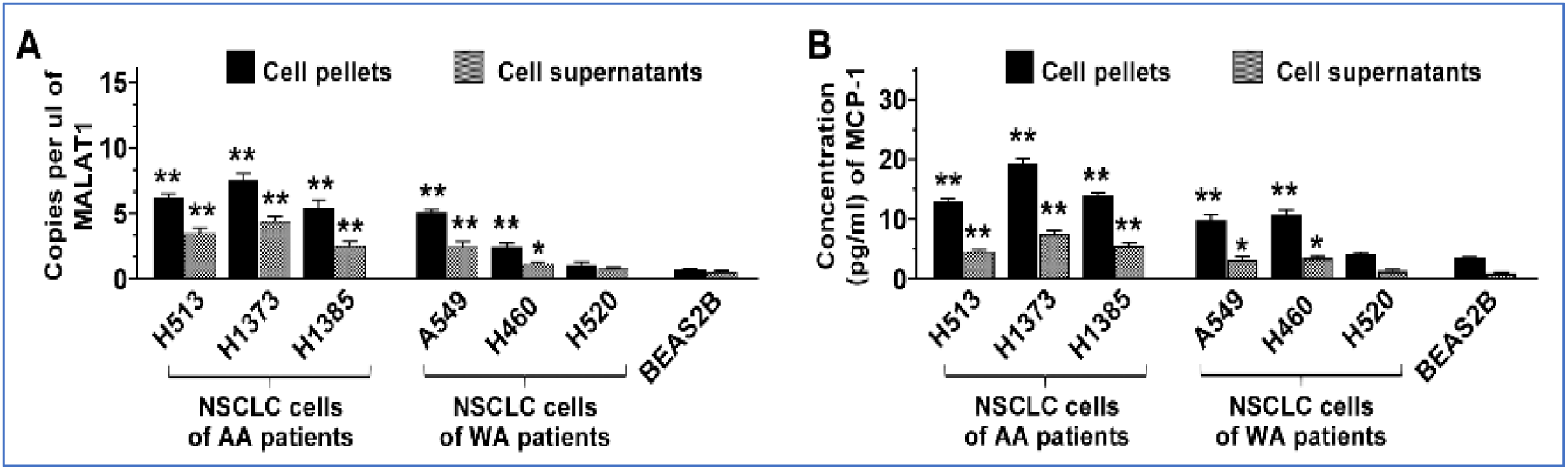
MALAT1 and MCP-1 are expressed in cancer cell lines. A. This bar chart shows the quantification of MALAT1 in NSCLC cell lines, with individual bars representing cellular pelle and supernatant samples. The MALAT1 levels in NSCLC cell lines, specifically in H513, H1373, A549, H460, and H1385, are elevated in both cells and supernatants compared to normal cells (BEAS2B), with significant differences observed (*p<0.05; **p<0.01). B. MCP-1 levels in NSCLC cel lines, particularly in H513, H1373, A549, H460, and H1385, are significantly higher in both cells and supernatants compared to normal cells (BEAS2B), indicating a notable increase (*p<0.05; **p<0.01)

**Fig. S2.**
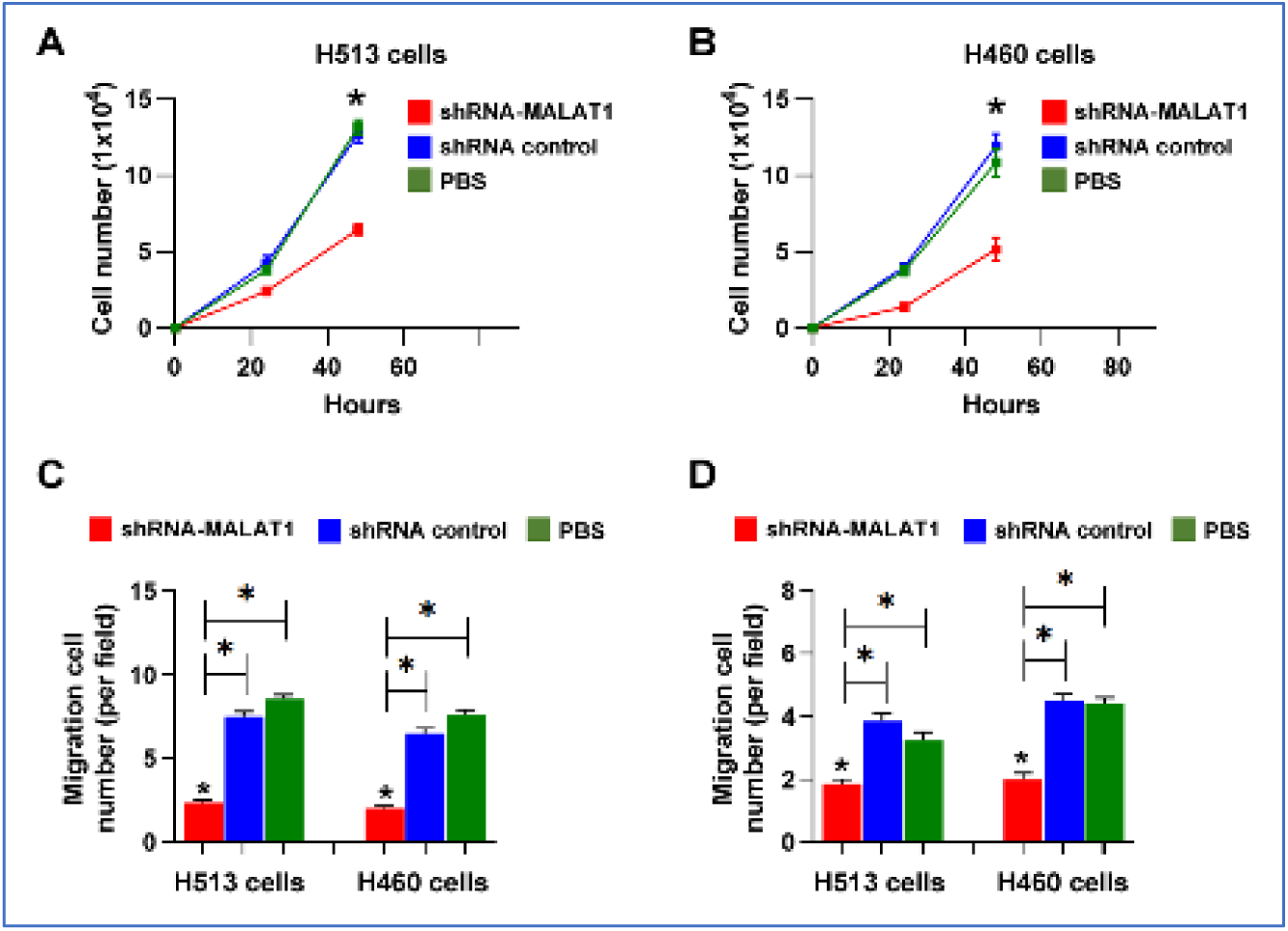
Effects of MALAT1 knockdown on the proliferation and migration of H513 and H460 cancer cells. A. Proliferation of H513 cells over time. The graph shows the cell number on the y-axis against time in hours on the x-axis. Three groups are compared: cells treated with shRNA-MALAT1, shRNA control, and PBS. The shRNA-MALAT1 treatment significantly reduced the proliferation rate compared to the control and PBS groups, as indicated by the lower cell counts at each time point. *, all p<0.05. B. Proliferation of H460 cells over time. Similar to panel A, cell numbers are plotted against time for the three groups: shRNA-MALAT1, shRNA control, and PBS. The shRNA-MALAT1 treatment group shows a marked decrease in cell proliferation in comparison to the control and PBS treatments. *, all p<0.05. C-D. Migration assay shows a significant reduction in cell migration compared to the shRNA control and PBS groups for both H513 and H460 cells. Error bars represent the standard deviation of the mean from three experiments. *, all p<0.05.

**Fig. S3.**
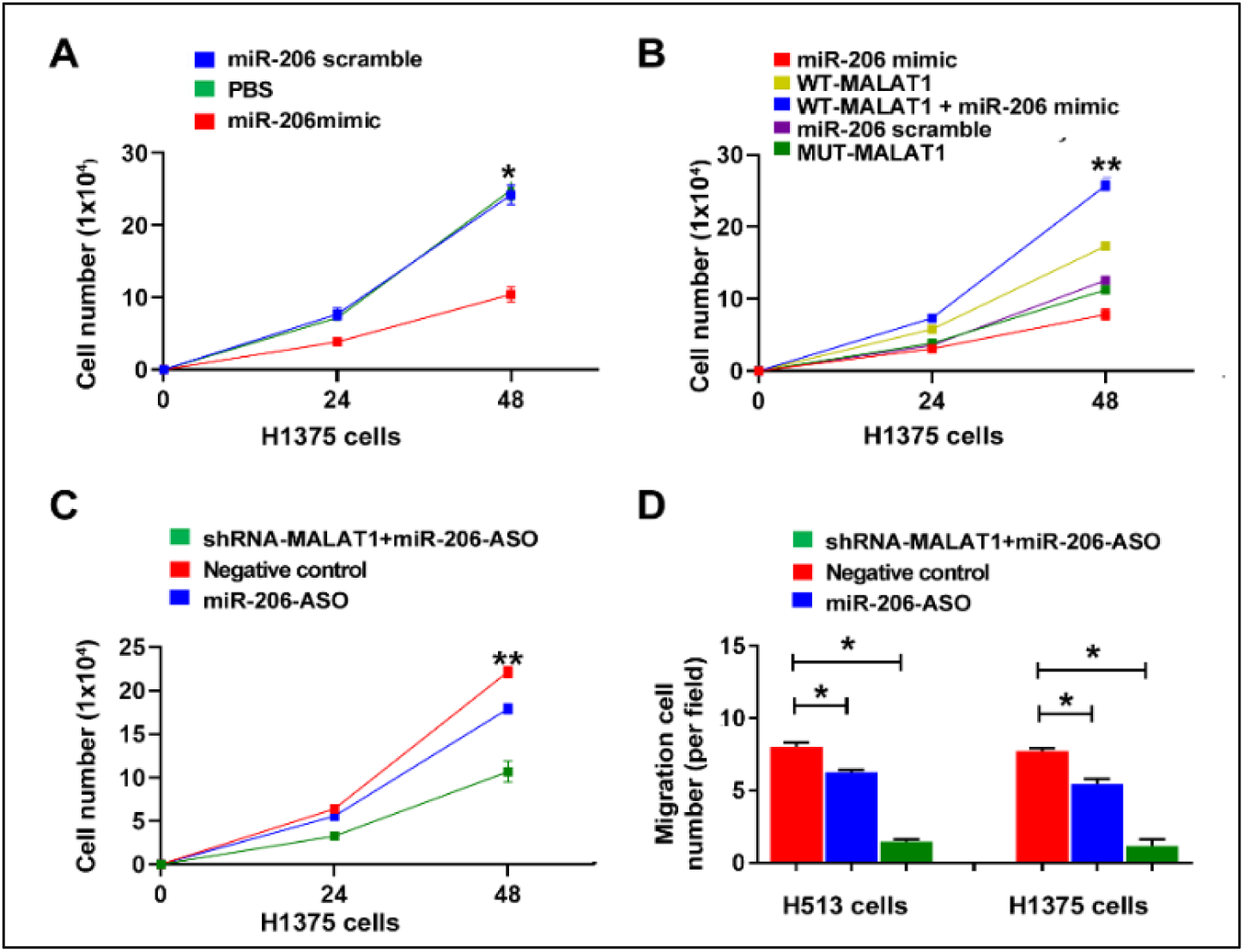
MALAT1 directly targets miR-206, influencing MCP-1 expression, and contributes to the lung tumorigenesis. A. Proliferation assay showing cell numbers of lung cancer cells (H1375) treated with either miR-206 mimic or miR-206 scramble control over a 48-hour period. *, p<0.01. B. Cell proliferation assays for H1385 cells treated with miR-206 mimic, WT-MALAT1, WT-MALAT1 plus miR-206 mimic, miR-206 scramble, or MUT-MALAT1 over 48 hours. *, all p<0.05; **, all p<0.01. C. Proliferation assay of H1375 cells treated with sh-MALAT1 plus miR-206-ASO, negative control, or miR-206-ASO alone, showing restored proliferation with miR-206 ASO treatment. *, all p<0.05; **, all p<0.01. D. Migration assay for H513 and H1375 cells demonstrates the reversion of sh-MALAT1 tumor-suppressive effects by miR-206 ASO. *, all p<0.05.

